# Clinical profiles at the time of diagnosis of COVID-19 in Costa Rica during the pre-vaccination period using a machine learning approach

**DOI:** 10.1101/2021.06.18.21259157

**Authors:** Jose Arturo Molina-Mora, Alejandra González, Sergio Jiménez-Morgan, Estela Cordero-Laurent, Hebleen Brenes, Claudio Soto-Garita, Jorge Sequeira-Soto, Francisco Duarte-Martínez

**Affiliations:** Centro de Investigación en Enfermedades Tropicales (CIET) & Facultad de Microbiología, Universidad de Costa Rica, San José, Costa Rica; Instituto Costarricense de Investigación y Enseñanza en Nutrición y Salud (INCIENSA), Tres Ríos, Costa Rica; Escuela de Medicina, Universidad de Costa Rica, San José, Costa Rica

**Keywords:** COVID-19, Costa Rica, machine learning, symptoms, time of diagnosis, SARS-CoV-2, clustering, clinical profiles

## Abstract

**Background:** The clinical manifestations of COVID-19 disease, caused by the SARS-CoV-2 virus, define a large spectrum of symptoms that are mainly dependent on the human host conditions. In Costa Rica, almost 319 000 cases have been reported during the first third of 2021, contrasting to the 590 000 fully vaccinated people. In the pre-vaccination period (the year 2020), this country accumulated 169 321 cases and 2185 deaths.

**Methods:** To describe the clinical presentations at the time of diagnosis of COVID-19 in Costa Rica during the pre-vaccination period, we implemented a symptom-based clustering using machine learning to identify clusters or clinical profiles among 18 974 records of positive cases. Profiles were compared based on symptoms, risk factors, viral load, and genomic features of the SARS-CoV-2 sequence.

**Results:** A total of seven COVID-19 clinical profiles were identified, which were characterized by a specific composition of symptoms. In the comparison between clusters, a lower viral load was found for the asymptomatic group, while the risk factors and the SARS-CoV-2 genomic features were distributed among all the clusters. No other distribution patterns were found for age, sex, vital status, and hospitalization.

**Conclusion:** During the pre-vaccination time in Costa Rica, the clinical manifestations at the time of diagnosis of COVID-19 were described in seven profiles. The host co-morbidities and the SARS-CoV-2 genotypes are not specific of a particular profile, rather they are present in all the groups, including asymptomatic cases. In further analyses, these results will be compared against the profiles of cases during the vaccination period.

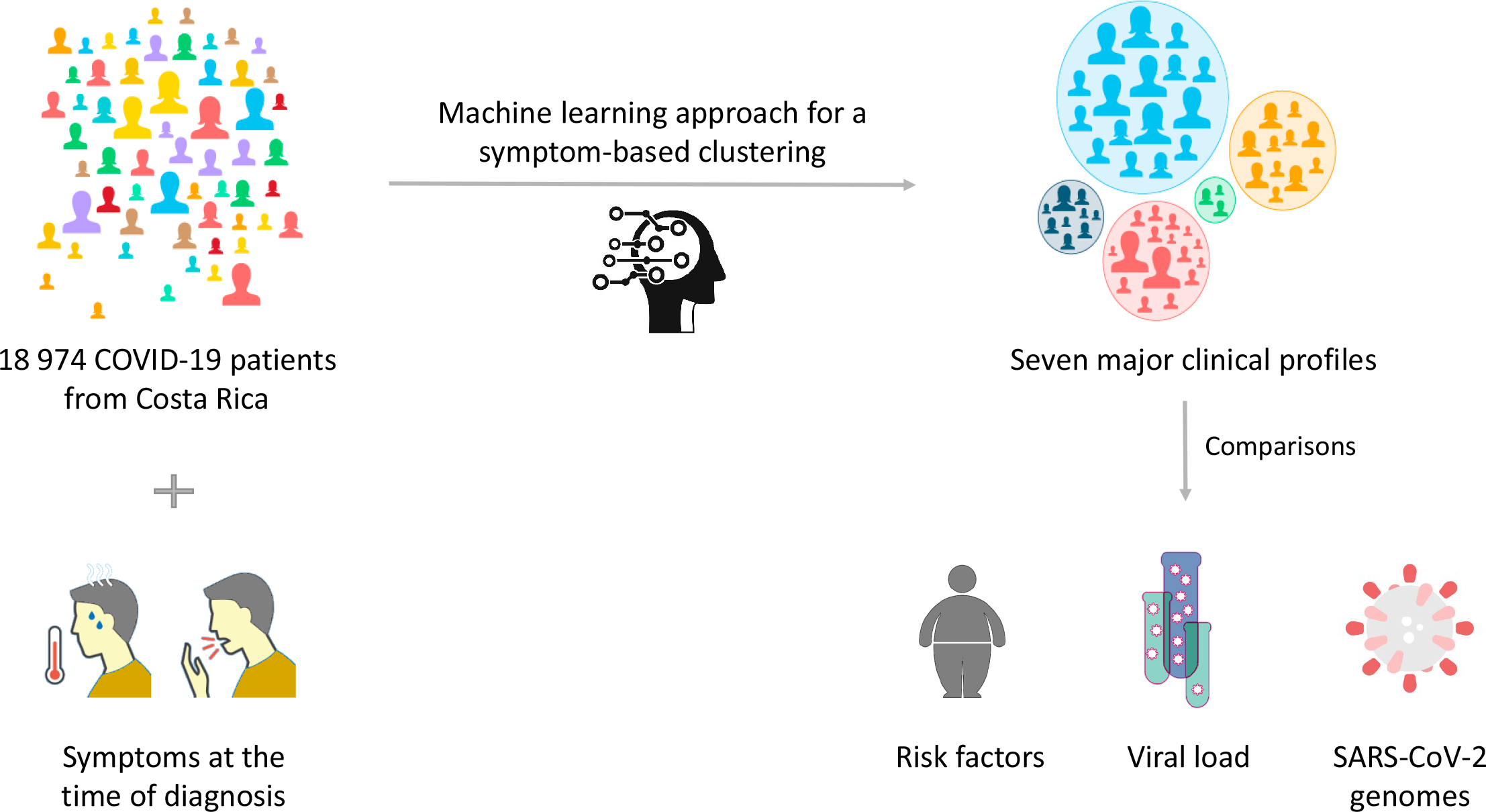

## Introduction

The COVID-19 (COronaVIrus Disease 2019) pandemic has impacted the public health systems around the world, even though a new hope was established with the beginning of the vaccination program at the end of 2020. In Costa Rica, until May 31^th^ 2021, almost 319 000 cases of COVID-19 and 2500 deaths have been reported (https://www.ministeriodesalud.go.cr/). On the same date, 590 000 people from Costa Rica (12% of the population) have been fully vaccinated after the first application was started on December 24^th^ 2020. In the pre-vaccination period (the year 2020), this country accumulated 169 321 COVID-19 cases and 2185 deaths.

As an infectious disease, the spread and manifestations of the infection by SARS-CoV-2 depend on the agent (the SARS-CoV-2 virus), the human host (comorbidities and genetic factors), and the environment (social interactions, containment measures, etc.) (Tsui, Deng, & Pan, 2020). In our previous work, we focused on the analysis of the genomic diversity of SARS-CoV-2 sequences from Costa Rica during 2020 (Molina-Mora et al., 2021) and we now studied the symptoms at the time of diagnosis of COVID-19 as clinical profiles.

The clinical manifestations of COVID-19 include asymptomatic cases or patients with a non-specific clinical presentation. Symptomatic cases report a variety of symptoms, including fever, anosmia, cough, and diarrhea; more severe cases are reported with respiratory distress, sepsis, septic shock, and death (Huang et al., 2020). Due to the diversity of symptoms, human factors such as genetics and risk factors play a critical role in the outcome of the disease (LoPresti, Beck, Duggal, Cummings, & Solomon, 2020; Sironi et al., 2020; Toyoshima, Nemoto, Matsumoto, Nakamura, & Kiyotani, 2020). These factors tend to be specific to the population, in which particular studies are required in each geographic location. In addition, many patients are evaluated only at the time of diagnosis due to the clinical presentation of a mild illness, in which the tracking of symptoms is lack or not possible later. This point-out the need for defining clinical profiles at the initial stages of the COVID-19, for example at the time of diagnosis. Besides, it is expectable that the beginning of the vaccination strategy affects the spread and the clinical manifestations of COVID-19 cases (Amit et al., 2021), which can be eventually contrasted with the pre-vaccination pandemic time.

On the other hand, the diversity and mainly the amount of COVID-19 patients define a complex challenge in the step of data analysis to describe the clinical features in the populations. To overcome this situation, clustering or unsupervised machine learning approaches bring an opportunity to extract relevant information by identifying patterns, clusters, or profiles within large volumes of data. Although some machine learning or similar approaches have been implemented to investigate clinical symptoms from patients with COVID-19 worldwide (Dixon et al., 2021; Fu et al., 2020; Han et al., 2020; Kim et al., 2020; Sudre et al., 2021; Tong, Wong, Zhu, Fastenberg, & Tham, 2020), to our knowledge, none has been formally reported from Costa Rican cases.

Therefore, because of the relevance of describing local clinical profiles in the early stages of COVID-19 disease in a pre-vaccination pandemic period, and the use of strategies to deal with massive data, this work aimed to identify and describe clinical profiles at the time of diagnosis of COVID-19 in Costa Rica during 2020 with a symptom-based clustering approach using machine learning.

## Methods

### Data source, software, and pre-processing

This is an observational retrospective study with COVID-19 patients from Costa Rica. Initially, 68 758 records of suspected patients were included. Data corresponded to all the registered cases in INCIENSA (Instituto Costarricense de Investigación y Enseñanza en Nutrición y Salud, the institution in charge of the epidemiological surveillance in Costa Rica) during the year 2020 (between March 6 and December 31, 2020).

All the different analyses for pre-processing, machine learning approaches, and visualization were performed with custom scripts in the RStudio software (Version 1.1.453, https://www.rstudio.com/) with the R software (Version 3.6.3, https://www.r-project.org/) in local servers of the Universidad de Costa Rica. The following packages were used during this implementation: “caret”, “haven”, “RColorBrewer”, “ggfortify”, “cluster”, “plotrix”, “ggpubr”, and “randomcoloR” (details in https://cran.r-project.org/web/packages/).

For the pre-processing step, different filtering, cleaning, and re-arrangement strategies were applied to data, as follows. We only considered cases with positive results by PCR test for SARS-CoV-2, without repeated tests (for patients with multiple tests, we only selected the first record), completing 18 974 records. Each record was composed of 121 epidemiological and clinical (symptoms at the time of diagnosis and risk factors) data and the viral load by the Ct value in the PCR assay. For 160 cases, genomic information of the viral sequences of SARS-CoV-2 (clade and lineage, and the presence of the mutation spike-T1117I of the Costa Rican lineage B.1.1.389) was available from our previous work (Molina-Mora et al., 2021), which was included for the comparisons.

### Clustering analysis by a machine learning approach

To identify major groups of COVID-19 patients based on the symptomatology at the time of diagnosis, a clustering analysis was completed with all the 18 974 records. Although there were 51 distinct symptoms among the patients, most of them were of very low frequency. Thus, we only included symptoms present in at least 1% of the patients, with a final selection of 18 symptoms (a small group of symptomatic patients with only “rare” or low-frequency symptoms was analyzed as non-symptomatic cases at this step).

Afterward, to define the groups based on the 18 symptoms of the 18 974 patients, a machine learning strategy was implemented using Hierarchical Clustering (HC). To select the best conditions for the clustering analysis, we followed three main steps. First, to define how different were the clinical manifestations among all patients, we assessed five different distance metrics (Euclidean, Binary, Maximum, Manhattan, and Minkowski). The optimal metric had to identify a separated group for the “asymptomatic cases”. Second, the Elbow criterion was implemented to determine the expected number of major clusters, by plotting the explained variation as a function of the number of clusters (Shi et al., 2021). The number of clusters K was defined according to the elbow of the curve and, due to this is a heuristic approach, a tolerance of 1 was considered (i.e., number of clusters = K±1). Finally, using the optimal distance metric and the expected number of clusters, the tree was cut using a single height value to define the clusters. Groups with at least 5% of the cases (949 out of the 18 974 patients) were labeled as major clusters, and the remaining small groups were included in a single “sink” cluster.

### Clusters comparison

After the definition of the major clusters, the groups were compared using demographic data (age, sex, localization, etc.), clinical information (symptoms, risk factors, vital status, hospitalization, Ct value, etc.), and SARS-CoV-2 genotypes (clades, lineages, and presence of the spike-T1117I mutation of the Costa Rican lineage B.1.1.389). To this end, representation of comparisons was done using heatmaps, barplots, and boxplots, with the subsequent statistical tests by ANOVA, Tukey test, Chi-square, and other tests as appropriate.

## Results

### Seven major clusters with specific symptoms define the clinical profiles at the time of diagnosis of COVID-19

In order to identify clinical profiles of COVID-19 patients based on 18 symptoms (present in at least 1% of the patients) at the time of diagnosis, we developed a clustering strategy using machine learning with 18 974 records. After data pre-processing, five distance metrics were assessed within the HC algorithm. The selection of the best metric was based on the ability to separate all the asymptomatic cases in a single group, which was only achieved when a Binary distance was implemented (Figure 1-A), unlike other approaches (Figure 1-B-C). To define the number of expected clusters, the Elbow criterion suggested k=8±1 as the optimal number (Figure 1-D).

**Figure 1.**
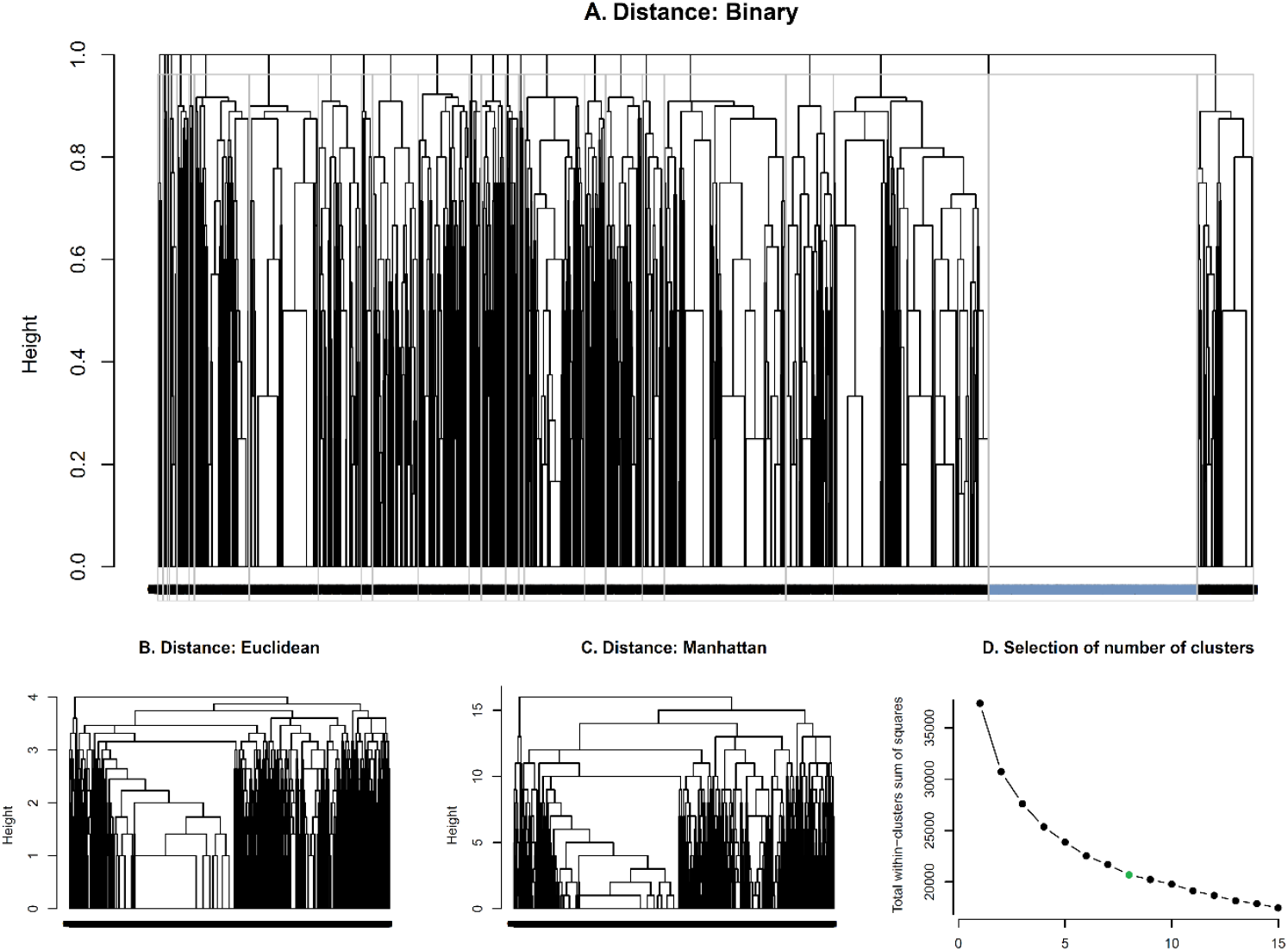
Parameters of the clustering using machine learning to identify clinical profiles of COVID-19 patients based on symptoms at the time of diagnosis. Using clinical data of 18 974 patients, different clustering analyses were run with different distance metrics, including Binary (A), Euclidean (B), and Manhattan (C). Only the Binary distance was able to cluster the asymptomatic cases in a single group, as expected (blue group). In the analysis using the Elbow criterion (D), the plot of variation identified the k=8 (green) as the number of expected clusters.

Using the parameters for the optimal clustering (distance and number of expected clusters), seven major clusters composed by at least 5% of cases (represented by non-gray colors) and a sink group (dark gray) were defined when the clustering tree was cut (Figure 2, top). See details of size for all the clusters in the Supplementary Material. The red cluster corresponded to the group with all the asymptomatic cases. As found in the heatmaps for all the patients (Figure 2) and the total frequency (Figure 3, left) the composition is dependent on the symptoms, as expected. See below for more details.

**Figure 2.**
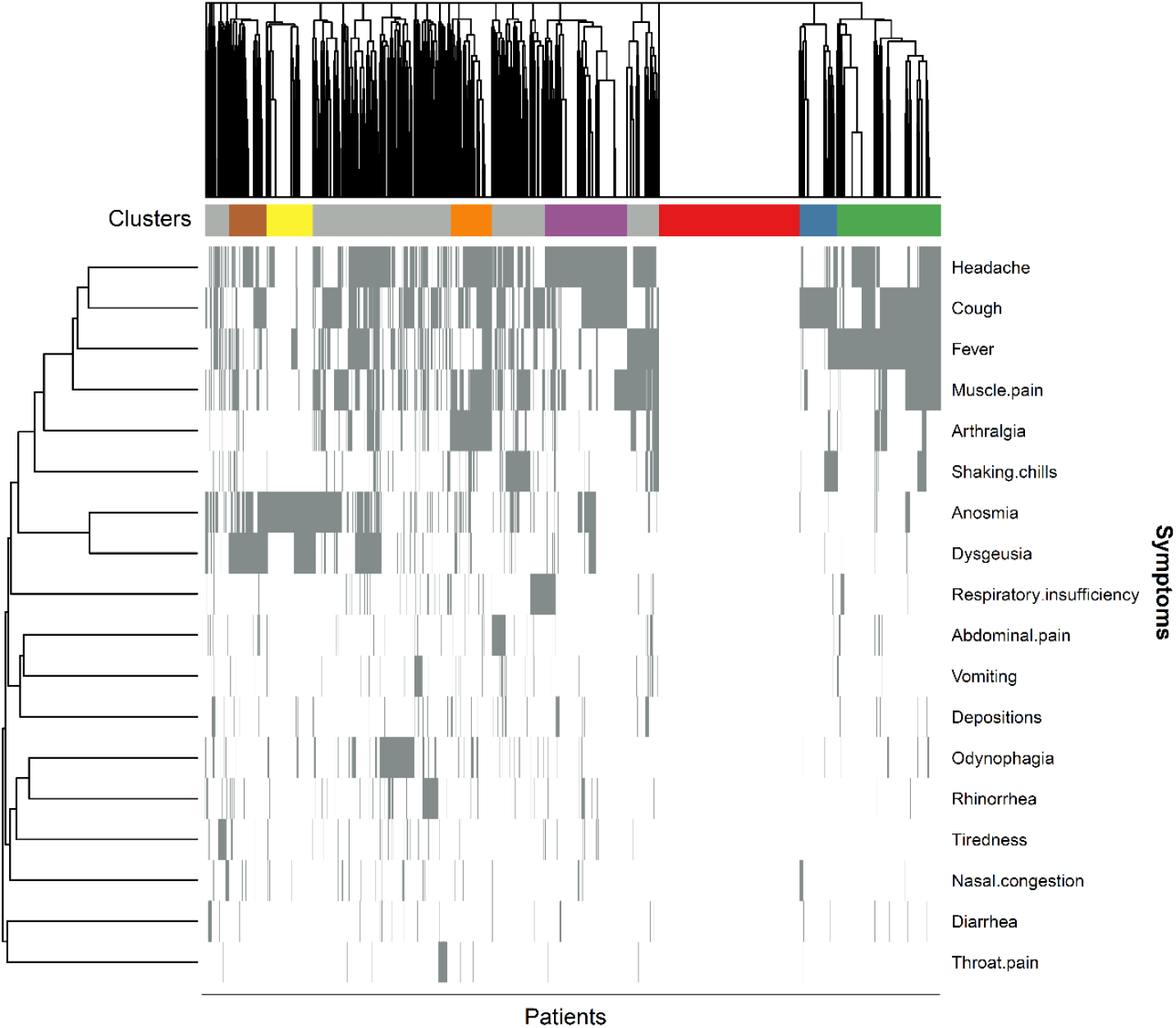
Seven major clinical profiles of COVID-19 patients were identified by a clustering approach using symptom information at the time of diagnosis. Seven major clusters (colors) and a sink group (dark gray) were defined, including a well-identified group for all the asymptomatic cases. Some symptoms co-occurred among patients (left dendrogram). In the heatmap, the presence or the absence of the symptom was represented by a light gray or white color, respectively.

**Figure 3.**
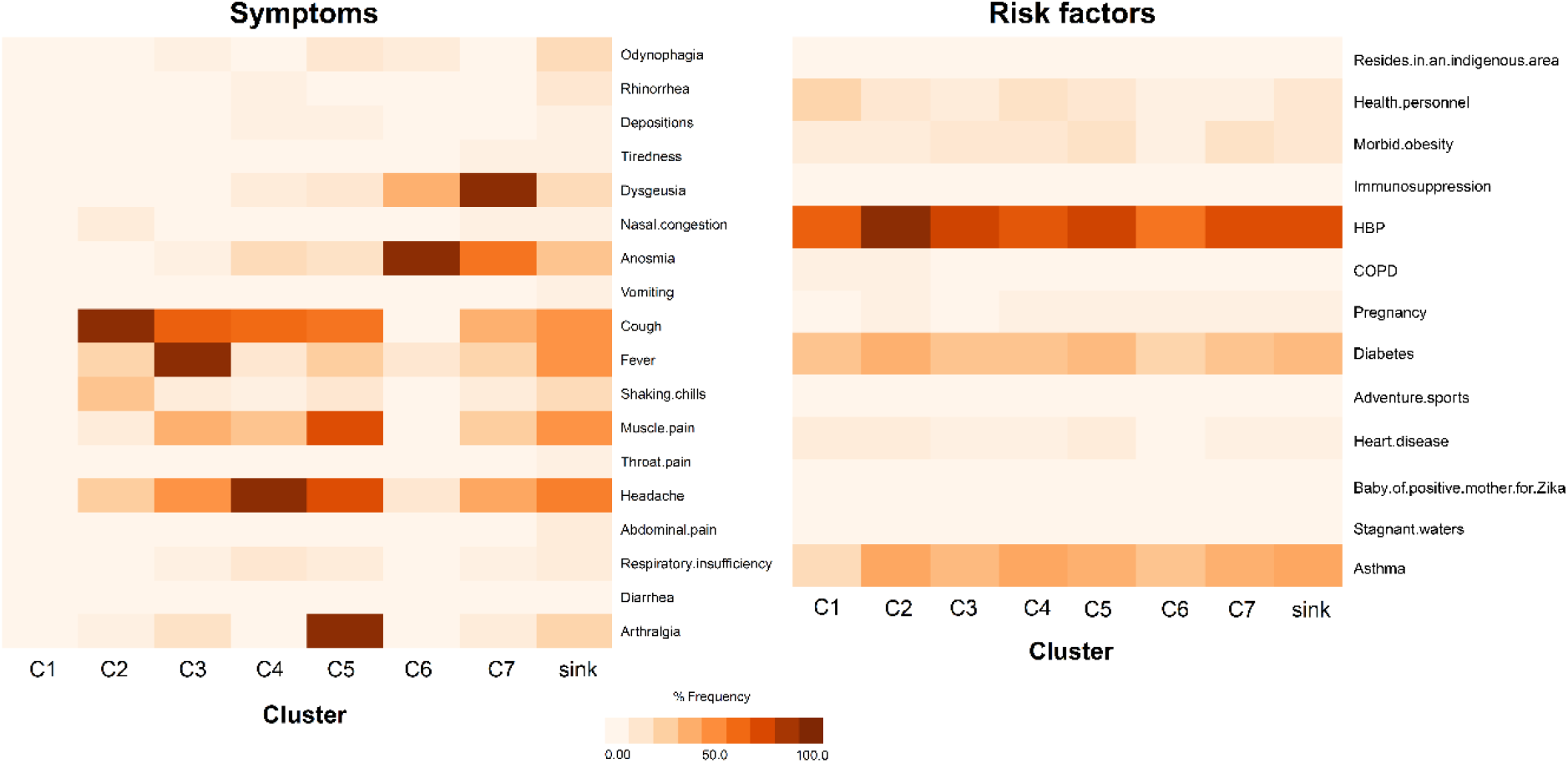
Frequency patterns of symptoms and risk factors of patients among the clusters of the clinical profile. Each cluster is composed of specific and predominant symptoms (left). The risk factors are distributed among all the clusters without any enriched pattern, including the asymptomatic and sink groups.

As shown in Table 1, major clusters are composed of between 953 and 3613 patients (also see Figure 4-A). The 3440 cases without any of the 18 main symptoms were found in cluster C1. The small fraction of 173 symptomatic cases in the C1 is the patients with “rare” or low-frequency symptoms (not included in the 18 used for the clustering), as expected. No other patterns regarding age, vital status, sex, nor hospitalization conditions were recognized, and these parameters were distributed similarly among clusters (Table 1, Figure 4-B and Supplementary Figure S1).

**Table 1.** Composition of clusters by epidemiological, clinical, and genomic data.

| Groups |  | Clusters |  |  |  |  |  |  |  |
| --- | --- | --- | --- | --- | --- | --- | --- | --- | --- |
|  |  | C1 | C2 | C3 | C4 | C5 | C6 | C7 | sink |
| Total patients |  | 3613 | 974 | 2683 | 2106 | 1042 | 1190 | 953 | 6409 |
| Sex | Female | 1776 | 460 | 1133 | 1119 | 510 | 616 | 506 | 3217 |
|  | Male | 1819 | 511 | 1541 | 979 | 529 | 572 | 444 | 3175 |
|  | ND | 18 | 3 | 9 | 8 | 3 | 2 | 3 | 17 |
| Symptoms | Yes | 173 | 974 | 2683 | 2106 | 1042 | 1190 | 953 | 6409 |
|  | No | 3440 | 0 | 0 | 0 | 0 | 0 | 0 | 0 |
| Hospitalized | Yes | 52 | 19 | 42 | 34 | 11 | 13 | 15 | 105 |
|  | No | 1260 | 304 | 716 | 529 | 220 | 353 | 243 | 1553 |
|  | ND | 2301 | 651 | 1925 | 1543 | 811 | 824 | 695 | 4751 |
| Alive (vital status) | Yes | 2840 | 792 | 2225 | 1772 | 896 | 973 | 830 | 5429 |
|  | No | 9 | 0 | 7 | 0 | 0 | 1 | 1 | 7 |
|  | ND | 764 | 182 | 451 | 334 | 146 | 216 | 122 | 973 |
| Number of distinct GISAID clades* |  | 3 | 4 | 3 | 3 | 3 | 1 | 2 | 4 |
| Number of PANGOLIN lineages* |  | 7 | 6 | 10 | 4 | 5 | 1 | 4 | 8 |
| Presence of the mutation spike-T1117I* | Yes | 11 | 5 | 7 | 7 | 3 | 2 | 1 | 14 |
|  | No | 34 | 8 | 30 | 12 | 4 | 0 | 3 | 19 |

\* Based on 160 genomes; ND: No Data

**Figure 4.**
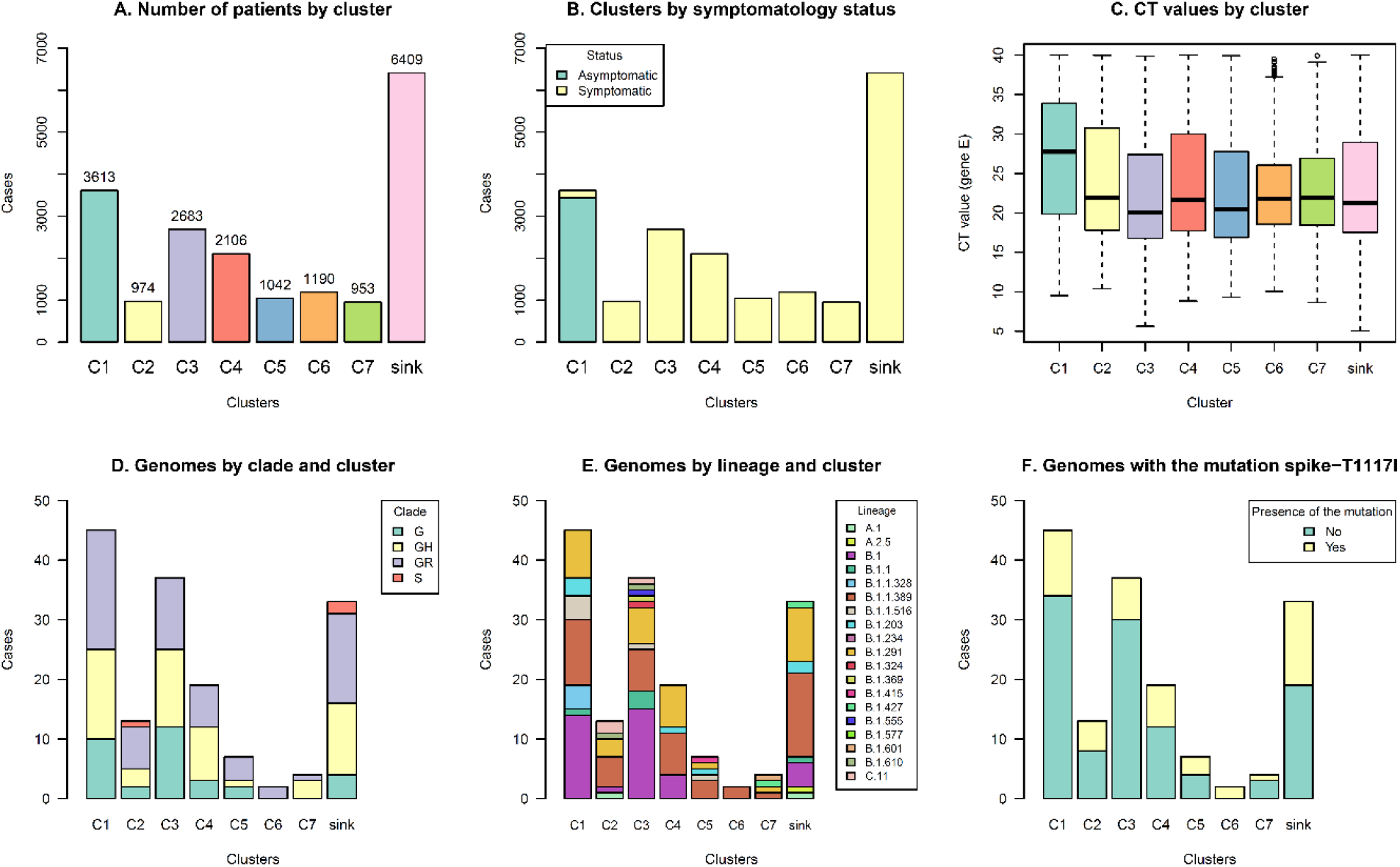
Distribution of demographic, clinical, and SARS-CoV-2 genomic information of cases of COVID-19 among seven major clusters. Major clusters are composed of 953-3613 patients. All the asymptomatic cases are found in the same cluster C1. Interestingly, the viral load (inferred from the Ct value) is lower in this group. Different SARS-CoV-2 genomes (lineages, clades, and the presence of the mutation T1117I in the spike) were distributed among all the clinical profiles.

Analysis of the co-presence of symptoms among the patients (Figure 2, columns), several symptoms were clustered (rows, left side). For example, there is a cluster of general symptoms (Figure 2, top left), digestive conditions (middle left), or more respiratory symptoms (down left).

In the comparison between symptoms (Figure 3, left), each cluster has a specific clinical profile. Cluster C1 is the group of all the asymptomatic cases. The C2 is characterized mainly by the presence of cough and rarely other symptoms. In contrast, C3 and C4 include cough and another main symptom (fever and headache, respectively). C5 is mainly composed of four symptoms, including arthralgia as the header. The conditions of anosmia and dysgeusia are the major components of the C6 and C7 clusters, with an inverted pattern of frequency.

### Risk factors and diverse SARS-CoV-2 genomes are distributed among all the clinical profiles, and viral load inferred from Ct values was lower for asymptomatic cases

Concerning the description of the risk factors among the clusters (Figure 3 right), all the conditions are present in all the groups without specific patterns, including the C1 for asymptomatic patients and the sink. The conditions with higher frequency are high blood pressure (HBP), asthma, and diabetes among all the profiles. Interestingly, asthma was found in a less frequency for the asymptomatic group, and HBP has a higher frequency in patients of cluster C2.

About the expected viral load (Figure 4-C), interestingly the Ct values for cluster C1 of asymptomatic cases were higher in comparison to all the other clusters (p<0.05). See statistical details in the Supplementary Material.

On the other hand, using 160 cases in which the SARS-CoV-2 genome was sequenced, it was possible to infer that the SARS-CoV-2 clades and lineages were not associated with specific symptoms nor clinical profiles, and they are distributed among all the clusters (Figure 4-D-E). This also applies to the Costa Rican lineage B.1.1.389 (orange in the barplots of Figure 4-E), which carries the mutation spike-T1117I and was the most common detected lineage during 2020 in the country (Figure 4-F), which is not specific to a particular profile.

## Discussion

The clinical manifestations of COVID-19 define a large spectrum of symptoms, as found in other studies (Fu et al., 2020; Kim et al., 2020; Sudre et al., 2021). Estimates of the features and proportion of the distinct clinical manifestations of COVID-19, including asymptomatic cases, are vital parameters for modeling studies (Byambasuren et al., 2020). In addition, early identification of symptoms is important for successful diagnosis, medical management, and treatment selection (Kostopoulou et al., 2015). This is a key point for health professionals that are in charge of gathering symptoms information when testing patients (the time of diagnosis during the first point of contact), to be able to differentiate between the most and least prevalent clinical presentation of COVID-19 in a specific community. In this regard, we studied the clinical profile at the time of diagnosis of 18 974 COVID-19 patients from Costa Rica during 2020 (the pre-vaccination period).

At the time of diagnosis, 18 symptoms were found to be present in at least 1% of the COVID-19 patients from Costa Rica, including non-specific symptoms (fever, headache, etc), as well as respiratory and gastrointestinal manifestations. Using a machine learning approach, seven major clusters or clinical profiles were found with those symptoms. The clusters showed the expected heterogeneity in the clinical presentation among COVID-19 patients from Costa Rica, just as it has been observed worldwide according to hundreds of case reports (Dixon et al., 2021; Fu et al., 2020; Han et al., 2020; Kim et al., 2020; Sudre et al., 2021; Tong et al., 2020). Besides, six main symptoms are defining the clinical profiles (Figure 3) and that must be taken into higher consideration at the moment of filling a patient’s chart: cough, fever, headache, arthralgia, anosmia, and dysgeusia. Congruently, most of these manifestations are included in the limited number of symptoms that are known to be associated with infectious diseases (Jeon, Baruah, Sarabadani, & Palanica, 2020). Also, the general description of the clinical manifestations can be used as part of the “case definition of COVID-19” given by the local and international epidemiological surveillance systems (World Health Organization, 2021).

A multivariable logistic regression and exploratory factor analysis by (Dixon et al., 2021) determined five symptom clusters among which ageusia, anosmia, and fever tend to be highly associated with SARS-CoV-2 infection, which resembles our findings in cluster C6. This also supports other findings in a meta-analysis in which up to 52.73% and 43.93% of COVID-19 patients presented olfactory and gustatory dysfunction, respectively (Tong et al., 2020), also found in cluster C7. In a second cluster, (Dixon et al., 2021) reported shortness of breath, cough, and chest pain, but only the cough had a high frequency in our data (cluster C2) without being associated with those other two symptoms. A third cluster was composed of fatigue, muscle ache, and headache. Of those symptoms, we only found headache as the main symptom in cluster C4. Finally, the last two clusters reported were represented by vomiting and diarrhea, and a runny nose with a sore throat (Dixon et al., 2021). None of those two clusters coincides with our findings. As Figure 3 shows, even if digestive symptoms are present among Costa Rican COVID-19 patients from C1 to C7, their frequency is very low. Nonetheless, this should not be neglected as it has been reported that some individuals present digestive symptoms alone, which is of clinical relevance as those patients may last longer achieving viral clearance compared to those with associated respiratory symptoms (Han et al., 2020).

In another work, a similar approach with machine learning techniques for the study of COVID-19 symptoms, six temporal profiles were identified after self-reported data were used (Sudre et al., 2021). To make a better comparison, day 0 symptoms were contrasted with our findings. Interestingly, dysgeusia was not included as the main symptom in their study, even though was the most prevalent one in our cluster C7. Cough and fever were found to be associated with the second cluster reported by (Sudre et al., 2021) as well as in profile C3 in our study. Headaches were distributed among all the clusters in both studies.

About risk factors, three chronic diseases were found among Costa Rican patients in all of the seven clusters. From most to least prevalent, the most significant conditions were high blood pressure, diabetes, and asthma. Interestingly, this finding is highly consistent with a meta-analysis by (Yang et al., 2020), who reported that the most prevalent comorbidities among SARS-CoV-2 patients were hypertension (21.1%), diabetes (9.7%), cardiovascular disease (8.4%), and respiratory system disease (1.5%). This is of clinical relevance to take these comorbidities into account when performing a screening among COVID-19 tests. However, we identified no reliance on the co-morbidities and the clinical profiles for COVID-19 patients. This result is in line with a meta-analysis that reported that up to 90% of clinical and demographic variables showed inconsistent associations with COVID-19 outcomes (Jeon et al., 2020).

Despite consulting several databases, no other works using machine learning were found using symptoms, risk factors nor SARS-CoV-2 genomic data of COVID-19 patients, and none using the initial clinical profile at the time of diagnosis. Machine learning techniques prove to be a very useful approach to study the variety of COVID-19 symptoms when large sets of data are available. The heterogeneity of this disease’s clinical presentation is reduced using this technique, thus it may help clinicians heighten vigilance of some specific symptoms over others.

On the other hand, the cluster of asymptomatic cases (C1) represents 18% of the total positive cases. This percentage is in line with other analysis in which the asymptomatic cases vary between 15 and 30% (Byambasuren et al., 2020; Centers for Disease Control and Prevention US, 2021), although other studies found higher frequencies (Byambasuren et al., 2020; Lee et al., 2020). The comparison of expected viral load between symptomatic and asymptomatic cases, using the Ct value, has been also reported as very variable (Trunfio et al., 2021; Tutuncu, Ozgur, & Karamese, 2021). Similar to our findings in which the symptomatic groups had lower Ct values, another study reported that higher viral load was associated with more signs and symptoms at diagnosis and a more frequent pattern of respiratory and systemic complaints (Trunfio et al., 2021). However, no associations between viral load and symptoms state have been also suggested in other works (Lee et al., 2020; Tutuncu et al., 2021). The situation of very diverse patterns of Ct values and clinical outcome is a drawback that can be explained not only by the individual factors, but also the technology, sample quality, and the time of sampling after infection (Buchan et al., 2020). Therefore, this complex scenario implies that there is not consensus between the initial viral load and the clinical manifestations of COVID-19 (Byambasuren et al., 2020; Trunfio et al., 2021).

Regarding the SARS-CoV-2 genotypes, our reports of the independence of the clinical presentation of COVID-19 and the genomic determinants of the SARS-CoV-2 sequence are in line with others studies (Grubaugh, Hanage, & Rasmussen, 2020; Hodcroft et al., 2020; van Dorp et al., 2020). For each cluster, a diversity of clades and lineages were identified, including independence of the presence or absence of the mutation T1117I from the Costa Rican lineage B.1.1.389 (Molina-Mora et al., 2021). This situation reminds us that the clinical profiles depend on the viral agent and human host conditions. The human genetic, comorbidities, and risk conditions have been described as the predominant factor in the clinical outcome of the COVID-19 disease, as found in several studies (LoPresti et al., 2020; Molina-Mora et al., 2021; Sironi et al., 2020; Toyoshima et al., 2020).

Furthermore, owing to the distribution of SARS-CoV-2 genotypes among all the clusters, our results suggest that genomic features of the virus are not associated with specific changes in the clinical presentation, as has been reported recently, including relevant variants (Graham et al., 2021; Nakamichi et al., 2021). The lack of change in symptoms for different SARS-CoV-2 genotypes also indicates that existing testing and surveillance infrastructure do not need to change specifically for these versions of the SARS-CoV-2 genome (Graham et al., 2021).

Our analyses presented some limitations that must be taken into account in the interpretation of results: (1) classification of positive cases of COVID-19 was based on the positivity of a PCR for nasopharyngeal samples, i.e., we depended on the performance of the test and sample quality; (2) records were retrieved from a local database with some missing information, mainly for SARS-CoV-2 genomic data; and (3) symptoms of very low frequency, social behavior, or genetic factors of the host were not considered in this study.

Finally, due to vaccination started massively in January 2021 in Costa Rica (although the first doses were applied at the end of December 2020), we consider that this study represents a special work to give the panorama of COVID-19 in pre-vaccination time (2020). In future work, we hope to assess the vaccination status and how this event has impacted the clinical profiles of COVID-19 patients during 2021.

## Conclusion

In conclusion, the identification of seven clinical profiles at the time of diagnosis of COVID-19 was achieved using a clustering approach. In general, 18 symptoms were reported in at least 1% of the COVID-19 patients from Costa Rica, although six clinical manifestations were predominant. A specific symptom frequency was revealed for each cluster or clinical profile. In the comparison between clusters, a higher viral load inferred from the Ct values was found for the asymptomatic group, while the risk factors and the SARS-CoV-2 genomic features were distributed among all the clusters. Therefore, the host co-morbidities and the SARS-CoV-2 genotypes are not specific of a particular profile, rather they are present in all the groups, including asymptomatic cases. No other distribution patterns were found for age, sex, vital status, and hospitalization.

Jointly, these results describe the clinical manifestations at the time of diagnosis of COVID-19 in Costa Rican patients during the pre-vaccination time of the pandemic, as well as they can be used for decision making by the local healthcare institutions (first point of contact with health professionals, case definition, infrastructure, etc). In further analyses, these clinical patterns will be compared against cases during the vaccination period.

## Data Availability

Processed data is found in the Supplementary material.

## Ethical approval and consent to participate

This study was approved by INCIENSA (INCIENSA-DG-of-2020-174) and the scientific committee of CIET-UCR (No. 242-2020). Data were collected for epidemiological surveillance according to the Costa Rican regulation Law Nº 8270 (May 17th, 2002), in which no additional consent was required for retrospective studies of archived and anonymized samples.

## Acknowledgments

We thank clinicians, microbiologists, and other personnel of the public (Caja Costarricense de Seguro Social CCSS) and private clinical laboratories for the samples of confirmed cases of COVID-19. We also thank members of CIET-Universidad de Costa Rica and INCIENSA for their logistic and financial support in the activities associated with the project.

## Availability of data and material

Processed data is found in the Supplementary material.

## Author contributions

J.M.M., H.B., C.S.G., and F.D.M. participated in the conception and design of the study. J.S.S. was responsible for data acquisition from INCIENSA database. J.M.M. and A.G. were involved in data pre-processing. J.M.M. implemented and standardized all the machine learning pipelines. J.M.M., S.J.M., E.C.L., H.B., C.S.G., J.S.S., and F.D.M. were involved in the interpretation of results. J.M.M. drafted the manuscript. All authors reviewed and approved the final manuscript.

## Figures and tables

**Figure S1.**
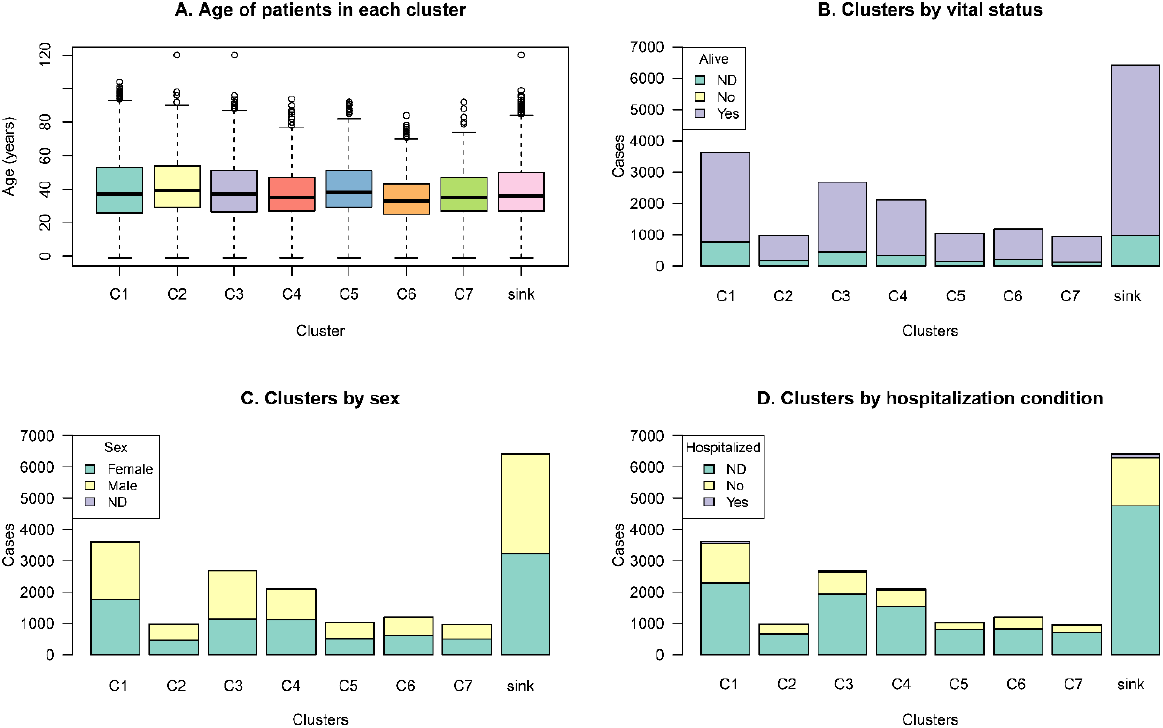
Distribution of patients among clusters according to age, vital status, sex, nor hospitalization conditions. No specific patterns are identified in the composition of the clusters.

